# Technology-Supported Self-Triage Decision Making: A Mixed-Methods Study

**DOI:** 10.1101/2024.09.12.24313558

**Authors:** Marvin Kopka, Sonja Mei Wang, Samira Kunz, Christine Schmid, Markus A. Feufel

**Affiliations:** Division of Ergonomics, Department of Psychology and Ergonomics (IPA), Technische Universität Berlin, Berlin, Germany; Department of Sociotechnical Systems, University of Wuppertal, Wuppertal, Germany

## Abstract

Symptom-Assessment Application (SAAs) and Large Language Models (LLMs) are increasingly used by laypeople to navigate care options. Although humans ultimately make a final decision when using these systems, previous research has typically examined the performance of humans and SAAs/LLMs separately. Thus, it is unclear how decision-making unfolds in such hybrid human-technology teams and if SAAs/LLMs can improve laypeople’s decisions. To address this gap, we conducted a convergent parallel mixed-methods study with semi-structured interviews and a randomized controlled trial. Our interview data revealed that in human-technology teams, decision-making is influenced by factors before, during, and after interaction. Users tend to rely on technology for information gathering and analysis but remain responsible for information integration and the final decision. Based on these results, we developed a model for technology-assisted self-triage decision-making. Our quantitative results indicate that when using a high-performing SAA, laypeople’s decision accuracy improved from 53.2% to 64.5% (OR = 2.52, p < .001). In contrast, decision accuracy remained unchanged when using a LLM (54.8% before vs. 54.2% after usage, p = .79). These findings highlight the importance of studying SAAs/LLMs with humans in the loop, as opposed to analyzing them in isolation.

## Introduction

Symptom-Assessment Applications (SAAs) are digital tools that help medical laypeople diagnose their symptoms and decide on appropriate care pathways^1^. Their primary goal is to empower individuals to make better decisions and to guide them to the most suitable care settings. On a systemic level this may ultimately alleviate pressure on overcrowded healthcare systems^2–6^. By optimizing care pathways, SAAs cannot only save time for patients but also free up vital resources for urgent healthcare needs. The financial costs of redirecting patients to more appropriate care settings is estimated to be more than $4 billion annually in the US^7–9^.

To achieve this goal, SAAs/LLMs must ensure that (1) the advice provided is accurate and (2) that it really improves users’ decisions. The accuracy of these applications has been extensively tested: Semigran et al. conducted a seminal study, which highlighted variability in performance – some symptom checkers were highly accurate while others performed poorly^5^. This initial study has been followed by numerous others that have replicated these findings, examined how accuracy evolves over time, and integrated real patient cases into their evaluations^3,10–14^. Further research has addressed methodological concerns to increase the validity and reliability of accuracy evaluation studies, such as disparities in testing procedures and the use of case vignettes that do not accurately reflect real-world scenarios^15–18^.

In addition to examining the accuracy of SAAs, several studies have explored human self-triage decision-making capabilities. For instance, Schmieding et al. discovered that on average, both SAAs and laypeople have a similar accuracy in making self-triage decisions, although the best-performing SAAs outperformed laypeople^19^. Levine et al. expanded this comparison to include the large language model (LLM) GPT-3 as an alternative to SAAs and found comparable accuracy levels between laypeople and GPT-3^20^. Additionally, Kopka et al. compared SAAs, LLMs, and laypeople and found that both SAAs and LLMs – although not all of them – achieved slightly higher accuracy in self-triage decisions than laypeople^18^. Thus, if the best performing SAAs and LLMs are chosen, they have the potential to improve laypeople’s self-triage decisions^18^.

However, users may choose to ignore even the best-performing SAAs/LLMs and thus not benefit from these systems. In other words, although users may outsource their decision-making partly and integrate the advice from SAAs/LLMs to varying extents, they ultimately make the final decision about which action to take. In a human-technology interaction context, two key concepts on human involvement can be distinguished: (1) human-out-of-the-loop, where the system operates entirely autonomously without human interference, and (2) human-in-the-loop in which humans make decisions and are actively involved^21^. For SAAs and LLMs used in self-triage, the setup is human-in-the-loop, as users ultimately make the final decision and take corresponding action. This concept has, however, not received much attention in SAA research yet. One study that used real-world data to assess patient care-seeking after using an SAA found that these tools might decrease perceived urgency among users^22^. However, in this study it was not possible to re-examine the cases to determine whether the decisions influenced by the SAAs were correct. Consequently, no conclusions can be drawn regarding performance. Another study indicated that many users tend to offload their decision-making heavily to SAAs, yet it also did not evaluate the performance of SAAs when being used by humans^23^. Thus, to empower individuals to make better self-triage decisions and reduce the burden on healthcare systems, it is essential to better understand laypeople’s self-triage decision-making with SAAs/LLMs in the loop and to evaluate the combined accuracy of SAAs/LLMs and laypeople.

As no model for understanding these self-triage decisions currently exists, we build on and adapt a stage model of diagnostic decision-making for physicians (see Figure 1) proposed by Kämmer et al., who used it to examine advice-taking in physician teams^24^.

**Figure 1.**
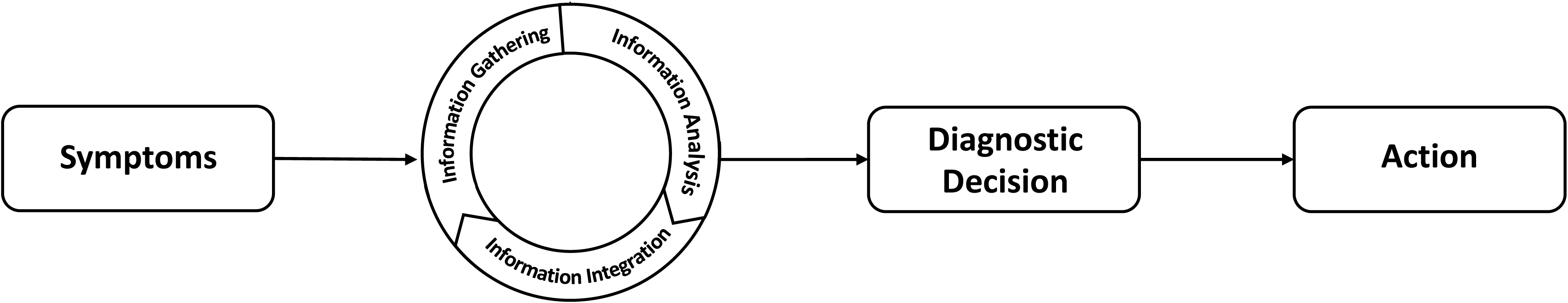
Stage Model of Diagnostic Decision-Making. Adapted from Kämmer et al. (2024)^24^ and the Committee on Diagnostic Error in Health Care (2015)^25^.

This model includes three main tasks: information gathering, information analysis, and information integration. When observing a patient’s symptoms, clinicians first seek to obtain more information about the symptoms and possible causes, then analyze this information to identify potential causes, and finally, integrate the collected evidence. This cycle may be repeated by gathering additional information after the initial decision, or it may conclude with a final decision that is subsequently implemented. Although the authors originally proposed this model for diagnostic decisions among medical professionals, it could also serve as a useful framework for examining self-triage decisions among medical laypeople.

In summary, it is currently unclear how users include the systems in their decision-making and if it makes their decisions more accurate. Based on prior research, decision-making might follow a similar path to diagnostic decisions among physicians and based on studies evaluating the accuracy of SAAs and LLMs in isolation, well-performing systems should enhance the self-triage decisions of laypeople. However, since these studies have not considered the human in the loop, it is unclear whether this translates to a better *‘human-SAA-team’* performance – which more closely resembles decisions in the real world – compared to a single human decision without SAA (or LLM) assistance. We aim to fill this research gap by addressing the following two research questions: How does the ‘human-in-the-loop’ team come to a decision? Do laypeople improve their self-triage decisions when using a well-performing SAA or LLM?

The first research question is exploratory and aims to understand processes within the human-SAA-team and their impact on decision-making. The second research question seeks to assess laypeople’s decision accuracy with SAAs/LLMs in the loop. Based on previous results from Kopka et al.^18^, we hypothesize the following:

H_1_: Laypeople have a higher self-triage accuracy with assistance of an SAA compared to making decisions on their own.

H_2_: Laypeople have a higher self-triage accuracy with assistance of an LLM compared to making decisions on their own.

## Methods

### Study Design

We used a convergent parallel mixed-methods design with semi-structured interviews to explore how humans integrate these systems into a human-in-the-loop decision and a Randomized Controlled Trial (RCT) to evaluate whether SAAs and LLMs can enhance the accuracy of participants’ self-triage decisions.

To gain insights into how SAAs and LLMs are used in users’ self-triage decision-making, we conducted narrative research and semi-structured interviews with participants before and after using these systems. These insights can help explore whether users merely adopt the recommendations they receive or if the decision-making process is more complex. This understanding could indicate whether isolated evaluation studies can predict real-world accuracy or if and why decisions from human-SAA teams deviate from the performance of SAAs alone.

To quantify the effect of using SAAs/LLMs in self-triage decisions, we conducted an RCT with a mixed parallel design (allocation ratio: 1:1). Participants were randomly assigned to receive advice either from ChatGPT or the symptom checker Ada Health (between-subjects factor) and assessed the appropriate self-triage level before and after receiving advice from the system (within-subjects factor). The allocation sequence was automatically generated using a simple randomization algorithm by the online survey tool SoSciSurvey, which also concealed the sequence by automatically assigning participants to their respective intervention groups. Due to the intervention’s nature, it was not possible to blind participants to their group assignment. The outcome assessor was blinded to group allocation until the statistical analysis was complete.

### Ethical Considerations

The ethics committee of the Department of Psychology and Ergonomics (IPA) at the Berlin Institute of Technology (tracking numbers AWB_KOP_3_230915 and 2225676) granted ethical approval for this study and informed consent was obtained from every participant prior to participating. The study was preregistered in the German Clinical Trials Register (DRKS00033775). The manuscript was constructed in accordance with the SRQR^26^ (for qualitative research) and the CONSORT^27^ (for RCTs) guidelines.

### Participants

The participants were required to be proficient in German and reside in Germany, as the questions and interviews pertained to specifics of the German healthcare system (e.g. the distinction between doctors in private practice and emergency medical services). Therefore, a certain degree of familiarity with the system was a prerequisite. They were excluded if they had professional medical training or participated in similar research from our department.

We recruited 24 participants for the interviews between November 2023 and January 2024 using a convenience sampling approach from our network and laid special emphasis on the diversity of participants to ensure different perspectives. Thus, we aimed to include 12 people with prior experience using digital triage tools (including telephone triage) and 12 people without prior experience. Participants were either family, friends, or part of the extended network of the interviewers. To accommodate varying schedules, interviews were conducted in the participant’s home, the interviewer’s home, or a university room, offering flexible settings intended to increase the comfort level and openness of the participants.

For the RCT, participants were sampled between the 14^th^ and 16^th^ March 2024 using a random sample from the sampling provider Prolific and were asked to fill out an online survey. To determine the required sample size, we conducted a simulation-based a-priori power analysis using R. Based on a previous study with a similar setup and the same vignettes^18^, we estimated laypeople’s accuracy at about 59%. Since the symptom checker Ada solved 19 out of 27 cases correctly, we estimated the accuracy to be about 67%. Using these values, we constructed a simulated dataset, specified a linear mixed-effects model (with a logit link function) and conducted the simulation-based power analysis with a desired power of 1-β = 0.80 and a significance level of α = .05. This resulted in a targeted sample size of 540 participants. Since we expected some users to answer inattentively, we oversampled by 10% and targeted a sample size of 600 participants. They received 0.53€ as a compensation for participation in the study, which took about 4 minutes. As a motivation to answer correctly and increase data quality, participants received an additional 0.06€ if they made the correct final decision^28^.

### Materials & Procedure

The interviews followed an interview guideline that was developed by SMW and SK, with refinements from all authors, and was pilot tested with 3 participants. Once finalized, the interview guide remained unchanged throughout data collection. Interviews were audio-recorded using smartphones. In the beginning, participants provided sociodemographic information. They were then randomized into using either the LLM ChatGPT or the SAA Ada. Participants then received one out of 27 validated case vignettes (stratified for each intervention group to ensure a similar distribution across both groups). The vignette set was taken from a previous study in which it was validated and constructed according to the RepVig Framework to ensure external validity^18,29^. The vignettes describe real cases from patients who (a) experienced the described symptoms, (b) prospectively wrote them while deciding if and where to seek care, and (c) consulted the internet for decision support. Thus, the vignettes have high generalizability to the use case SAAs are typically approached with. Participants were then asked how they would respond if they or someone close to them experienced these symptoms. They could choose from the following options, based on Levine et al.^20^: (1) ‘Call an ambulance or go directly to the emergency room. The symptoms must be treated immediately’, (2) ‘See or call a (family) doctor. The problem requires medical clarification, but is not a life-threatening emergency’, or (3) ‘The symptoms can be treated by yourself. It is probably not necessary to see a doctor’. Participants then used either ChatGPT or the SAA Ada to get a recommendation on what they should do. Throughout the interaction, they verbalized their thoughts in real-time using the think-aloud method^30^. In cases in which think-aloud did not work properly, participants were asked again to tell us what they were currently thinking. Following this interaction, participants reassessed the self-triage level of the case and explained their reasoning for the final decision made. The interviews were transcribed verbatim using MAXQDA with an adapted GAT 2 system.

In the RCT, participants followed the same approach: they were asked about their sociodemographic characteristics, previous experience with SAAs and LLMs, their health using the Minimum European Health Module^31^, their self-efficacy using the Allgemeine Selbstwirksamkeit Kurzskala^32^ and their technological affinity using the Affinity for Technology Scale Short^33^. Afterwards, they were presented with the case vignette and asked how they would respond if they or someone close to them experienced these symptoms. Participants were then randomized into one out of two intervention groups: receiving advice from either the LLM ChatGPT or from the SAA Ada. Given the nature of the intervention, they knew which system they saw and were not blinded. Participants were shown the result of ChatGPT (obtained using the prompt “Dear ChatGPT, I have a medical problem and hope you can give me some advice. The following are my symptoms: [Case Vignette] How urgent do you think it is and do I need to see a doctor or take other action?”, which represents a synthesis of how participants in the qualitative part asked their questions) or of the SAA Ada, which was selected because it was one the best-performing SAAs in previous studies^10,18^. We decided to show the corresponding results screens rather than direct interaction with the systems to maintain internal validity. After viewing the results, participants reassessed the self-triage level they thought was most appropriate. The primary outcome was self-triage accuracy and the secondary outcome was the change in urgency.

### Data Analysis

For the qualitative analysis, we applied reflexive thematic analysis as outlined by Braun & Clarke^34^ using MAXQDA. We chose this method and an inductive approach due to its suitability for identifying emergent themes in studies where complex decision-making processes are examined without predefined hypotheses^34^. The analysis began with an initial familiarization with the data, followed by the generation of initial codes. These were grouped into categories and themes, which were iteratively revised with the whole research team until they reflected the data and addressed the research question.

The quantitative data were analyzed using the symptomcheckR package, which is designed for analyzing self-triage data^35^. We used a mixed-effects regression model with the participant as a random effect and both the time point and system as fixed effects. The primary outcome, accuracy, was analyzed using binomial logistic regression. The secondary outcome, participants’ perceived urgency, was analyzed descriptively. To identify differences between the systems for the primary outcome, we conducted contrast tests that controlled for participants’ initial decisions. The p-values were adjusted for multiple comparisons by controlling the false discovery rate using the Benjamini-Hochberg procedure^36^.

## Results

### Participants

In the interviews, 24 people participated. Their characteristics are shown in Table 1. In the quantitative study, 631 people started the survey, of whom 16 were excluded because they were medical professionals and 10 who did not finish the questionnaire. Subsequently, we excluded two people because they failed the attention check (before randomization), one because they did not read the vignette (in the ChatGPT group), one because of self-reported technical problems (in the SAA group) and one for stating that their data should be excluded for other reasons (in the SAA group). Thus, the final dataset included data from 600 participants (301 in the SAA group and 209 in the ChatGPT group). Their characteristics are shown in Table 2.

**Table 1.**
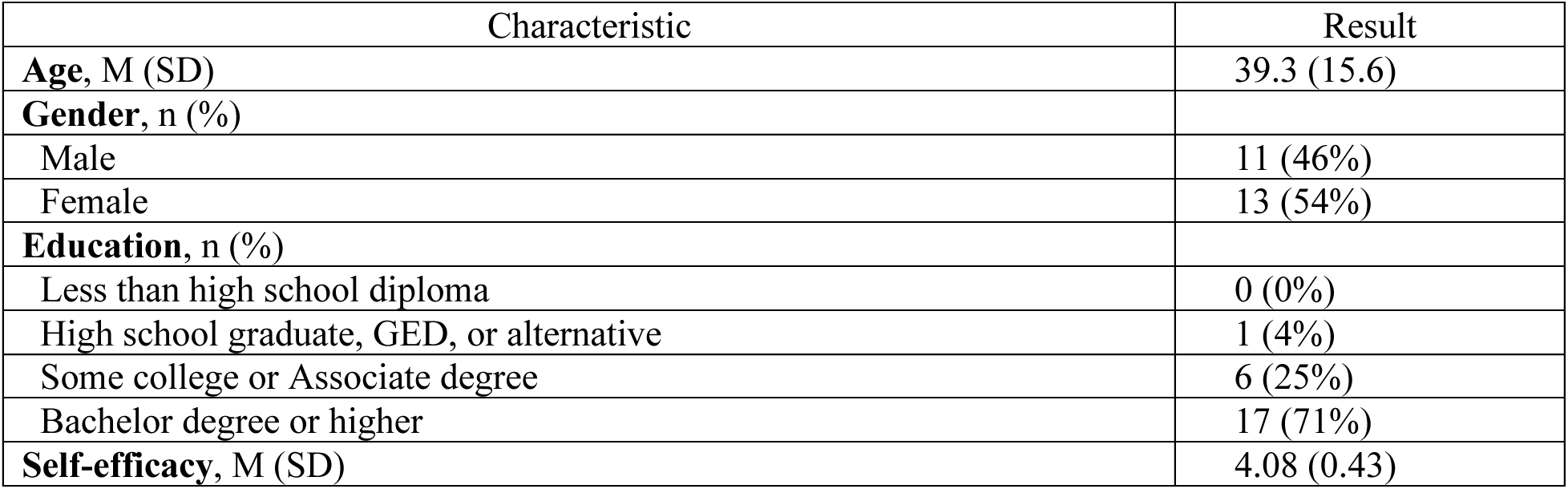
Description of participants in the interviews. N = 24, M = Mean, SD = Standard Deviation, n = number.

| Characteristic | Result |
| --- | --- |
| **Age, M (SD)** | 39.3 (15.6) |
| **Gender, n (%)** |  |
| Male | 11 (46%) |
| Female | 13 (54%) |
| **Education, n (%)** |  |
| Less than high school diploma | 0 (0%) |
| High school graduate, GED, or alternative | 1 (4%) |
| Some college or Associate degree | 6 (25%) |
| Bachelor degree or higher | 17 (71%) |
| **Self-efficacy, M (SD)** | 4.08 (0.43) |

**Table 2.** Description of participants in the RCT. N = 600, M = Mean, SD = Standard Deviation, n = number.

| Characteristic | SAA Group (n = 301) | ChatGPT Group (n = 299) |
| --- | --- | --- |
| **Age, M (SD)** | 31.4 (9.97) | 30.6 (9.07) |
| **Gender, n (%)** |  |  |
| Male | 145 (48%) | 155 (52%) |
| Female | 128 (43%) | 118 (39%) |
| Diverse or not disclosed | 28 (9%) | 26 (9%) |
| **Education, n (%)** |  |  |
| Less than high school diploma | 15 (5%) | 15 (5%) |
| High school graduate, GED, or alternative | 93 (31%) | 79 (26%) |
| Some college or Associate degree | 32 (11%) | 38 (13%) |
| Bachelor degree | 74 (25%) | 83 (28%) |
| Graduate degree or higher | 87 (29%) | 84 (28%) |
| **Medical training, n (%)** |  |  |
| No training at all | 200 (66%) | 183 (61%) |
| Basic first aid training | 101 (34%) | 116 (39%) |
| **Previous experience with SAAs, n (%)** |  |  |
| Never used before | 228 (76%) | 223 (75%) |
| Tested them, but did not use them again | 51 (17%) | 54 (18%) |
| Use them from time to time | 20 (7%) | 20 (7%) |
| Use them regularly | 2 (<1%) | 2 (<1%) |
| **Previous experience with LLMs, n (%)** |  |  |
| Never used before | 57 (19%) | 43 (14%) |
| Tested them, but did not use them again | 64 (21%) | 59 (20%) |
| Use them from time to time | 116 (39%) | 110 (37%) |
| Use them regularly | 64 (21%) | 87 (29%) |
| **Self-efficacy, M (SD)** | 3.33 (0.46) | 3.39 (0.41) |
| **Affinity for technology, M (SD)** | 4.11 (1.03) | 4.15 (1.02) |

### Insights into Team Decision-Making

#### Identified Themes

We identified three themes that relate to decisional influences ‘before the interaction’, ‘during interaction’, and ‘after interaction’ with eight categories: (1) Certainty in own appraisal, (2) Expectations, (3) Data basis, (4) Perceived personalization, (5) Information gathering and information analysis, (6) Explainability, (7) Information integration, and (8) Difficulties in information integration. These themes and a corresponding summary are shown in Table 3.

**Table 3.** Identified themes and categories.

| Theme | Category | Subcategory |
| --- | --- | --- |
| Before interaction | Certainty in own assessment | Informational need as prerequisite for information gathering |
|  |  | Previous experience leads to direct decision |
|  | Expectations | General mistrust |
|  |  | Fear of misdiagnosis |
|  |  | System is perceived as superior to oneself (due to lack of own medical knowledge) |
|  | Data basis | SAA perceived as trustworthy because of medical knowledge base |
|  |  | Concerns over data basis of ChatGPT |
| During interaction | Information gathering and information analysis | Gathering information: get more information on symptoms |
|  |  | Analyzing information: narrow down decision-making space |
|  | Perceived personalization | Criticism of generic responses |
|  |  | Increased trust and reliance through perceived personalization |
|  |  | Feeling of social interaction increased trust |
|  | Explainability | Need for uncertainty quantification |
|  |  | Lack of explainability in both systems |
| After interaction | Information integration | Accepting system's recommendation because it fits knowledge or experience |
|  |  | Seeking additional information to verify recommendations |
|  | Difficulties in information integration | Too much information felt overwhelming |
|  |  | Contradicting recommendations lead to uncertainty and higher urgency |

#### Before Interaction

Before interacting with the system, participants expressed different levels of certainty in their own symptom assessments, which influenced whether they sought additional information. Participants entered the interaction with varying – both positive and negative – expectations and mentioned the data basis as a specific factor that influenced their expectations.

#### Certainty in Own Assessment

Participants often had an initial idea of whether symptoms require care. If they were very confident in their assessment, they decided directly and were unlikely to consult additional sources of information. However, if they were unsure about the symptoms or uncertain in their own assessment, they often sought more information:

> *Then I just think to myself: ‘from a common sense point of view, it is probably water retention if it happens often and she’s not in pain. (…) I would then not consult the internet and no AI either.* (P16)
>
> *If I had [these symptoms] now, I would google it – put symptoms together or think of something else so that I can rule out certain things. Or get a hint: Please go to the doctor.* (P13)

Participants frequently based their initial decision on personal experiences or those of others. If they had experienced similar symptoms before and decided on a course of action, they relied on that experience as a heuristic to make a similar decision:

> *I stay with my first thought (…). Probably because I have had it myself and I have just taken this past experience and imposed it on the person or I just relied on my past experience for this decision.* (P14)

Also based on prior experiences, though related to SAAs/LLMs rather than symptoms, were participants’ expectations about the tools’ performance before using them.

#### Expectations

Participants had both negative and positive expectations about the tools before their interaction. A concern that participants expressed was fear of misdiagnosis and general mistrust in SAAs/LLMs. For example, one participant stated:

> *I think a big problem is that misdiagnoses can lead to major psychological stress. Or even in the case of misdiagnoses that say there is nothing wrong, it can of course have a negative impact on health.* (P17)

Conversely, some participants had a high initial trust in the systems and strongly believed that the system is more knowledgeable than themselves. This was often attributed to their lack of medical expertise:

> *I would think that ChatGPT has more background knowledge than I do (…) and could therefore answer the question better if in doubt.* (P24)

This quote also points to a key factor influencing participants’ initial trust and expectations of the systems: how they perceive the system’s database in relation to their own knowledge.

#### Data Basis

Participants highlighted the data basis of both systems. For the SAA, they assumed that – because it was an approved medical device – the data basis must be reliable. In contrast, they viewed the data basis of ChatGPT as unclear, untrustworthy, and thus did not consider ChatGPT reliable:

> *Because I know it’s a database that doesn’t lie. There are causes and symptoms that are linked and there’s just (…) a combination of multiple causes or multiple symptoms that cause (…) certain conditions. And these models don’t lie if they’re fed with the right data.* (P13, used the SAA)
>
> *I think you would have to design a special machine learning model or something or link it to a database of medical facts, because as it is now, I don’t think it would bring sufficient plausibility or verifiability.* (P5, used ChatGPT)

Whereas the previous themes refer to aspects that emerged before participants used SAAs or LLMs, the following quotes describe aspects during their use.

#### During Interaction

When interacting with the system, participants needed a high degree of personalization to trust it. They used the system not only to gather more information but also to narrow down their decision options and to analyze the available information. Additionally, a high level of explainability helped them make informed decisions and assess the uncertainty of a specific course of action.

#### Information Gathering and Information Analysis

When using the SAA and ChatGPT, participants used the tools both to explore symptoms and get more information, and to analyze information to get closer to a solution. Those who felt less informed before, wanted to find information they had not previously considered before. Conversely, participants who felt well-informed based on their existing knowledge skipped this step. Both groups used the tools to analyze the information, reduce the decision space, and to ultimately get closer to a decision.

For example, a participant used it to obtain more information and said:

> *Well, the app as a tool is quite influential. Especially because you get a lot of information again, about things you have not considered before.* (P21)

Another participant used it to analyze the symptoms and reported the following:

> *So it’s really like a selection, simply that you’re given certain things and then (…) the choices are narrowed down and then you get closer and closer to the diagnosis.* (P3)

While this theme focuses on the content of the responses, the following two themes – perceived personalization and explainability – relate more to the form and format of the responses provided.

#### Perceived Personalization

Participants expressed a clear desire for personalized results from the symptom assessment process. They showed higher trust and were more likely to rely on the recommendations when they were personalized with respect to the specific situation described by the users. They neglected information from the system in instances where the system gave unspecific answers or overlooked information they had entered:

> *I honestly don’t feel like it advises me particularly well because the answer is very generic. So for example, the first sentence is: ‘it’s advisable to make a doctor’s appointment as soon as possible, especially if your symptoms are new, unexpected, (…) or worsening’. That’s the kind of answer you would write in a guide. But I already described my symptoms in the beginning. So (…) I would expect the program to skip the general instructions and respond personally to what I have written.* (P10, used ChatGPT)
>
> *It didn’t just respond superficially, but it also went a bit into detail from the description I gave, which I though was good. (…) Just always this going back to what I said: it’s been like that for months; it was a lump. Yes, this can mean different things with different implications, so all of this was trustworthy.* (P5, used ChatGPT)

The systems’ capabilities to provide personalized responses and – in the case of ChatGPT – hold social conversations that feel close to human contact led to high trust and made the system convincing:

> *This direct approach to the specific question, so not just this keyword search ‘And here are 50 suggestions that could be an answer’, but you get a direct, personal, trustworthy answer, as if you were talking to a real person. And that’s what creates this trust, this direct chat.* (P24)

#### Explainability

Participants found it helpful when the SAA provided quantifiable estimates of uncertainty alongside its recommendations, such as stating “x out of 10”. Conversely, they noted that ChatGPT did not offer any quantifiable uncertainty, which they found less helpful:

> *And then on the fifth place, ‘lateral malleolar fracture’. 4 out of 100 people. Oh wow. Well then, I’ll go with the most likely* (P6, used the SAA)
>
> *Just the statistics. So that’s missing. Well, I say, the diagnoses that ChatGPT throws out are very intensive and not very quantified. So, I’ll also say he throws around technical terms without knowing who he’s talking to (…) or how seriously I take it or how many people have actually got these diseases.* (P24, used ChatGPT)

Both systems were criticized for their lack of explainability, as participants would have wished to understand how the systems arrived at their recommendations:

> *I don’t feel like it’s explained enough here (…) how ChatGPT arrives at something else. So, the explanation for a specific recommendation is not presented. It’s not rule-based enough for me, let’s put it that way. It doesn’t say: ‘okay, it’s this [symptom], so I would say with a greater probability this [disease], because this was like that in the past as a result of this and that’.* (P11)

#### After Interaction

After completing the interaction, participants attempted to integrate the new information and recommendations they received. Based on their prior experience, expectations, and knowledge, they evaluated the recommendations and either accepted them, combined them with their own understanding, or sought additional information and thereby started another iteration of gathering and analyzing information. If participants faced difficulties in information integration, they relied on heuristics to quickly make a decision and often concluded that the situation was urgent.

#### Information Integration

Participants tried to integrate the information they received and critically appraised the recommendations. They generally did so by verifying the advice with their own information and previous experience. If the recommendation was easily integrable into their previous knowledge, they readily accepted the system’s recommendation:

> *I think the self-care measures that are presented are good and would be enough for me. I would also follow them, also because I can say in comparison to past experiences that similar things have helped and it works, so in my head it makes sense.* (P4)

During the information integration, some participants still felt an informational need and tried to cross-verify the recommendation with other sources of information. Therefore, they started the cycle of gathering and analyzing information again:

> *And yet, I would still seek out a few more sources of information. In a similar way, where, if I go to a doctor and get a diagnosis, I usually come home and then read up a bit more about it. (…) With ChatGPT, it might be more about checking if what it told me is correct.* (P4)

#### Difficulties in Information Integration

In some instances, participants faced difficulties integrating information and instead relied on heuristics to make a decision. If the information was overwhelming, participants found it challenging to integrate and instead sought care quickly to reduce this conflict:

> *[I would advise to] See a doctor very quickly (…) because there could be so many different diagnoses, which you wouldn’t think of before, I would simply, yes, see a doctor, because I can’t diagnose it myself at all. I have no idea whatsoever and before I go crazy, I would see a doctor as soon as possible.* (P3)

If the advice drastically contradicted their prior beliefs, integrating it with their existing knowledge was difficult. As a result, they opted for a more urgent level of care:

> *Because I feel so confused now? Because initially, before I consulted ChatGPT, I was pretty sure about my decision that it wasn’t urgent. And (…) I assumed that it was not urgent and somehow these answers have confused me now because of course he first listed the worst [diagnoses], so to speak, which I would somehow still rule out now.* (P16)
>
> *They are somehow very different things. (…) Now that I see it, I would go to the doctor really quickly because it could be different things and serious things.* (P5)

These qualitative and exploratory findings suggest that translating advice from SAAs/LLMs into action is not a linear process. Improving the accuracy of these tools does not necessarily result in a medically correct action, as participants often rely on heuristics, such as comparing the advice to their own past experiences.

### Decision Improvement

#### Change in Accuracy

In the RCT, participants increased their self-triage accuracy from 53.2% to 64.5% when using the SAA (OR = 2.52 [1.50 – 3.55], z = 3.75 p < .001) but did not show such an increase when using ChatGPT (54.8% pre vs. 54.2% post usage, z = –0.27, p = .79). The difference in accuracy when using the SAA versus ChatGPT was statistically significant (OR = 2.24 [1.50 – 3.89], z = 3.59, p < .001). Participants’ accuracy in detecting emergencies (SAA: 63.6% pre vs. 81.8% post, z = 1.34, p = .25; ChatGPT: 68.2% pre vs. 90.9% post, z = 1.72, p = .13) and non-emergency cases did not increase statistically significantly with either system (SAA: 82.8% pre vs. 83.4% post, z = 0.27, p = .79; ChatGPT: 83.8% pre vs. 85.7% post, z = 0.92, p = .45). However, they detected more self-care cases correctly when using the SAA (13.1% pre vs. 36.9% post, OR= 8.59 [3.47 – 14.2], z = 3.80, p < .001) and fewer when using ChatGPT (16.3% pre vs. 8.1% post, OR = 0.00005 [0.0000003 – 0.000002], z = –5.28, p < .001). These differences are shown in Figure 2.

**Figure 2.**
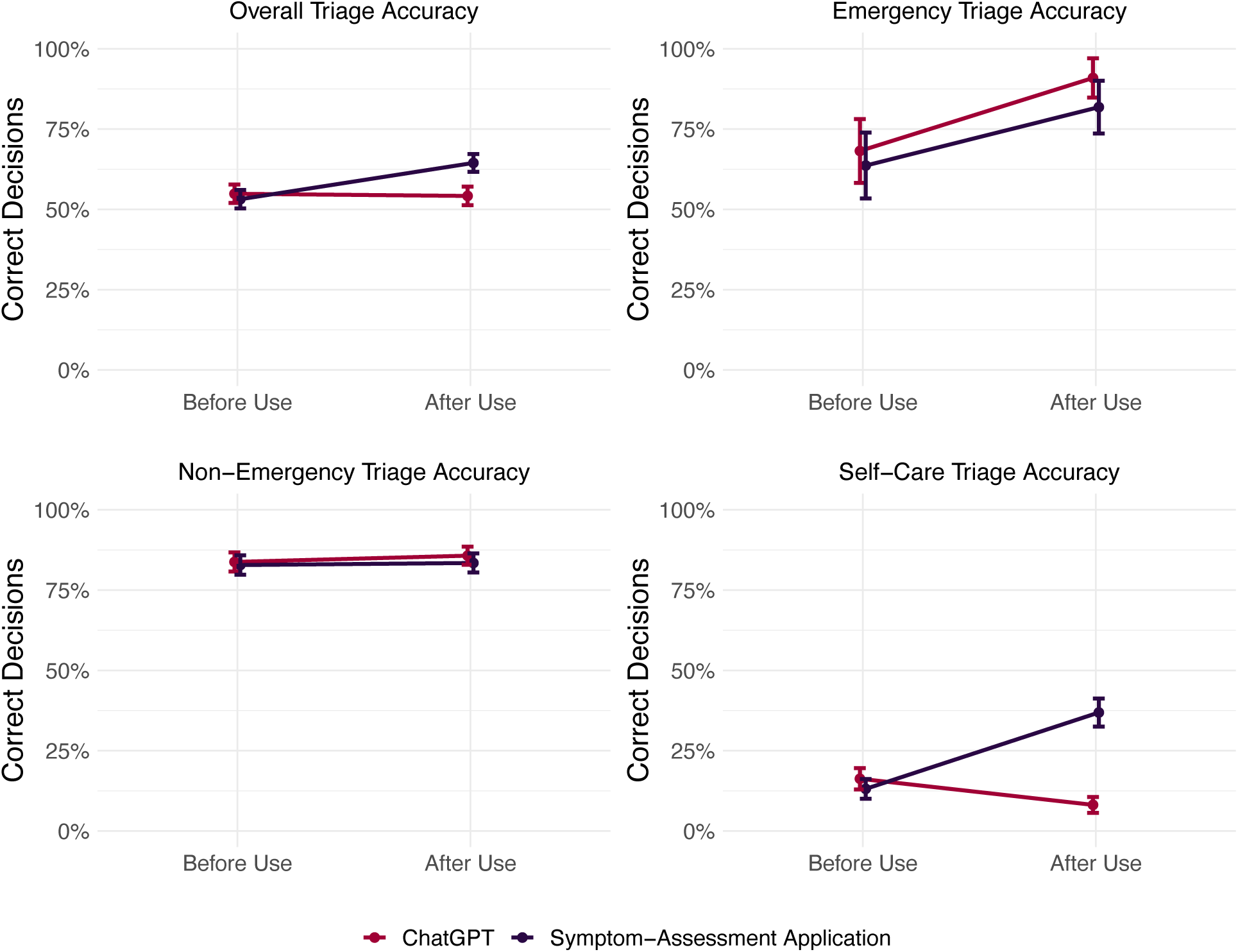
Change in self-triage accuracy when using the SAA Ada and ChatGPT.

In cases in which participants were initially correct but received incorrect advice from the SAA, they remained at a correct solution in 72% of cases (21/29). Conversely, if they were incorrect but received correct advice, they changed their appraisal to a correct decision in 37% (63/170) of all cases. The same was observed for ChatGPT: in cases in which participants were correct but received incorrect advice, they remained at the correct solution in 61% (22/36) of all cases. If they were incorrect but received correct advice, they changed their appraisal to a correct decision in 57% (38/52) of all cases.

#### Change in Urgency

Among participants seeing results from the SAA, most participants remained at their initial appraisal (73%, 221/301). If they changed it, 16% (48/301) decreased their urgency, whereas 11% (32/301) increased it. Among participants seeing results from ChatGPT, most participants remained at their initial appraisal as well (83%, 249/299). However, if they changed it, most participants increased their urgency (13%, 39/299) and only a minority decreased it (4%, 11/299). Urgency changes are shown in Figure 3.

**Figure 3.**
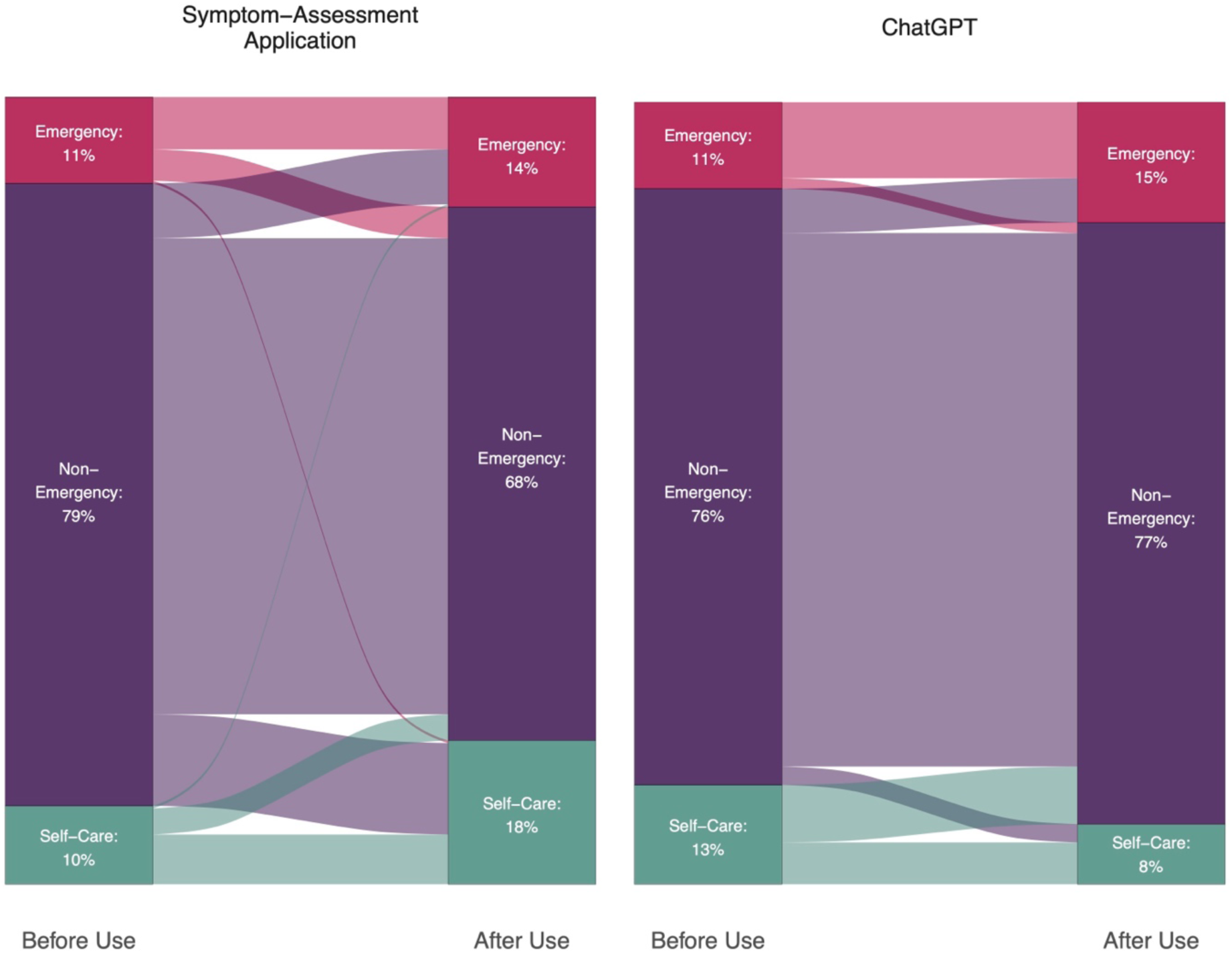
Change in urgency when using the SAA Ada and ChatGPT.

## Discussion

Our quantitative results demonstrate that laypeople make more accurate decisions when using a well-performing SAA compared to making decisions on their own, thus supporting our first hypothesis. However, this improvement was not observed when participants used ChatGPT for advice, leading us to reject our second hypothesis. Although laypeople did not achieve the high accuracy levels of the tested SAA, they approached these levels. Notably, they frequently ignored incorrect recommendations, especially when using ChatGPT. This observation suggests that while users benefit from correct advice, they are less frequently misled by incorrect suggestions. On a broader scale – since most interaction with SAAs in the real world refer to non-emergency or self-care cases^37^ and laypeople tend to be very risk-averse when unassisted^38^ – our findings indicate that using self-triage decision support systems like SAAs leads to a shift towards lower urgency. This result aligns with previous research, which highlighted the usefulness of SAAs in encouraging users to treat their conditions at home (when appropriate) and reducing unnecessary visits to healthcare facilities^22,39,40^. The results also demonstrate the importance of considering humans in the loop when evaluating SAAs and LLMs, as the performance of the team is different from isolated SAA accuracy.

Our qualitative part identified factors before, during, and after the interaction that influence the decision-making process in a *human-SAA team*. Before the interaction, participants’ certainty in their own assessments determined whether they sought additional information. If they did, their expectations and the system’s data basis influenced if and how they accepted advice from SAAs and LLMs. During the interaction, personalization and explainability played important roles: If users perceived a high degree of perceived personalization – particularly with LLMs as conversational agents – they had increased trust and were more likely to rely on the recommendations. Conversely, quantifying uncertainty seemed to decrease user’s reliance. These findings align with a systematic review on advice-taking^41^, which suggests that decision makers prefer advisors with high (perceived) expertise and relatability, which may be demonstrated in both systems. In the case of ChatGPT, not communicating uncertainty might have given the impression of high certainty and writing like a human being might have increased relatability by being perceived as a social agent. For the SAA, its use of highly professionalized language and the provision of extensive information may have led to being perceived as an expert system.

Our qualitative results can also be understood in relation to the stage model of diagnostic decision-making^24^. Factors before the interaction correspond to physicians’ existing knowledge used during the information gathering phase; factors during the interaction align with information gathering and analysis; and factors after the interaction correspond to information integration. From this perspective, users follow an approach similar to physicians: Their search for information is comparable to physicians’ inductive foraging (seeking more information on symptoms^42^) and their information analysis is similar to deductive inquiry (narrowing down the decision-space to approach a decision^43^). This corresponds to information gathering and information analysis in the stage model of diagnostic decision-making and users appear to use SAAs and LLMs specifically for these steps. Afterward – in line with the model – they integrate this information^24^. At this stage, most users critically evaluate the recommendation before making a final decision. This indicates that SAAs are often used to complement rather than replace individual decision-making.

Our findings can be used to adapt the stage-model of diagnostic decision-making^24^ for application in self-triage decisions. The resulting stage model of technology-assisted self-triage decision-making can be found in Figure 5.

**Figure 5.**
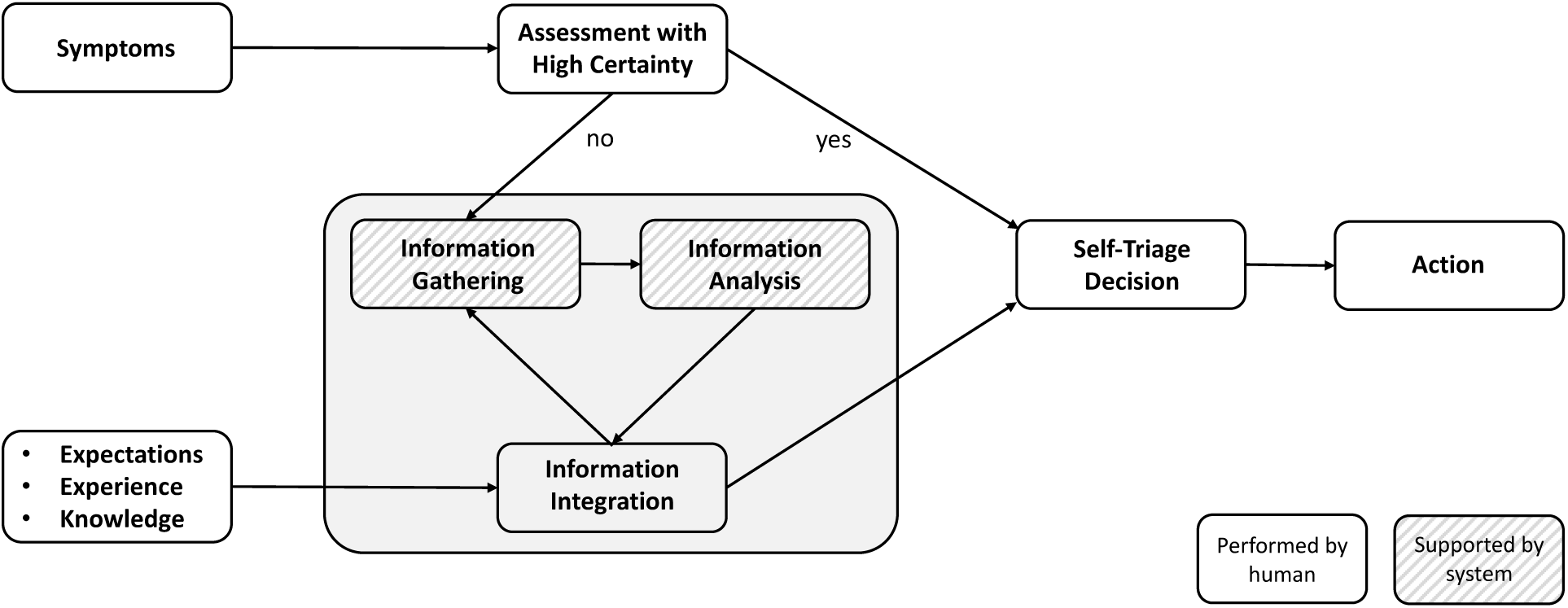
Stage model of technology-assisted self-triage decision-making.

The decision-making process begins when laypeople experience symptoms themselves or try to advise others. If they encountered similar symptoms before or are highly confident in an initial assessment of the appropriate care pathway, they make a decision directly based on their previous experience and knowledge. However, if they are uncertain about their initial assessment or have no idea what to do, they face an informational need and seek additional information, e.g., from technological systems^44^. They input their symptoms into the system (information gathering) which analyzes the information to provide a recommendation (information analysis). During the information integration phase, users try to integrate the recommendation and any other new information with their prior experience, expectations, and existing knowledge. If users believe they have identified the correct solution, they may conclude the process and determine a final self-triage level. In cases in which the recommendation and information is compatible with their previous information, they simply accept the received recommendation. Conversely, if the recommendation conflicts with their previous information, they weigh all informational cues to arrive at a decision. If they still face an informational need, they gather additional information and restart the cycle of information gathering and information analysis. However, if users encounter difficulties in information integration – such as feeling overwhelmed by too much information or facing highly conflicting information – they may abort the decision-making process and opt for a care pathway with high urgency. This approach allows them to reduce perceived uncertainty by deferring the integration of the wealth of complex information to a medical professional^45^.

In contrast to Kämmer et al.’s original model, our model suggests that users may not even engage in the full decision-making cycle. Instead, they might use a recognition heuristic to make a decision quickly^46^: They recognize the symptoms from previous experiences and directly choose the course of action that was successful in those situations. Additionally, our model differs from the original model in the allocation of tasks: In Kämmer et al.’s study^24^, information analysis and integration were important collaborative tasks in physician pairs, while information gathering was not a relevant part of the process^47^. In contrast, for teams comprising a layperson and an SAA, both information gathering and analysis seem to be allocated towards the SAA once it was consulted, whereas information integration is left solely to the human. The layperson thus typically has the role of a supervisor, makes the final decision and is thereby solely responsible for the information integration phase^48^.

This study is not without limitations. Although we used a validated set of case vignettes with greater external validity than traditional vignettes^18,29^, the nature of vignette-based studies still poses limitations compared to real-world evidence. Participants did not experience the symptoms themselves but only read about others’ symptoms, which could alter decision-making processes compared to experiencing symptoms directly. Nevertheless, SAAs are frequently used for other people as well^5^. Another limitation concerns our experimental setup. Whereas participants entered the symptoms themselves in the interviews, in the RCT they were entered by the study team and participants only saw the results screen. Although this setup limits generalizability, it increases internal validity by allowing us to examine the decision-making processes and the impacts of SAAs and LLMs in a controlled environment. However, future studies should replicate our results with higher external validity by allowing participants to input symptoms themselves. The experimental setup also guided our qualitative data analysis, which identified factors before, during, and after the interaction. Although this process is highly likely in real-world scenarios, there might be some instances where it is disrupted by external factors – for example, when outside individuals interrupt the decision-making process to provide their own input. Finally, we selected one of the best-performing SAAs for this study, as prior research suggests that only high-performing SAAs should be implemented^10,18^. Thus, it remains unclear how decisions might be influenced by a low-performing SAA and whether participants could counteract incorrect recommendations if they make up the majority of received recommendations.

In conclusion, laypeople seem to use SAAs and LLMs as decision aids rather than replacements. When working alongside SAAs as a human-SAA team, they make more accurate decisions than they would on their own, especially because they are able to compensate for incorrect recommendations. Given laypeople’s risk-averse nature, SAAs can be particularly effective in the real world to help identifying self-care cases correctly – a decision in which they outperform laypeople’s independent decisions. However, self-triage decision-making is complex, and the user’s role involves integrating all available information with their previous experience and knowledge to arrive at a decision. Thus, our findings highlight the importance of studying SAAs and laypeople not in isolation, but as integrated human-SAA teams where humans play an active role in the decision-making process.

## Data availability

The data will be published in an open access repositorium upon acceptance.

## Author contributions

MK and MAF conceived of the study. MK, SMW and SK collected the data. MK conducted the analyses and data visualization and wrote the first draft of the manuscript. All authors provided critical input and worked on manuscript development.

